# Towards Out-of-Lab Anterior Cruciate Ligament Injury Prevention and Rehabilitation Assessment: A Review of Portable Sensing Approaches

**DOI:** 10.1101/2022.10.19.22281252

**Authors:** Tian Tan, Anthony A. Gatti, Bingfei Fan, Kevin G. Shea, Seth L. Sherman, Scott D. Uhlrich, Jennifer L. Hicks, Scott L. Delp, Peter B. Shull, Akshay S. Chaudhari

## Abstract

Anterior cruciate ligament (ACL) injury and ACL reconstruction (ACLR) surgery are common. Many ACL-injured subjects develop osteoarthritis within a decade of injury, a major cause of disability without cure. Laboratory-based biomechanical assessment can evaluate ACL injury risk and rehabilitation progress after ACLR; however, lab-based measurements are expensive and inaccessible to a majority of people. Portable sensors such as wearables and cameras can be deployed during sporting activities, in clinics, and in patient homes for biomechanical assessment. Although many portable sensing approaches have demonstrated promising results during various assessments related to ACL injury, they have not yet been widely adopted as tools for ACL injury prevention training, evaluation of ACL reconstructions, and return-to-sport decision making. The purpose of this review is to summarize research on out-of-lab portable sensing applied to ACL and ACLR and offer our perspectives on new opportunities for future research and development. We identified 49 original research articles on out-of-lab ACL-related assessment; the most common sensing modalities were inertial measurement units (IMUs), depth cameras, and RGB cameras. The studies combined portable sensors with direct feature extraction, physics-based modeling, or machine learning to estimate a range of biomechanical parameters (e.g., knee kinematics and kinetics) during jump-landing tasks, cutting, squats, and gait. Many of the reviewed studies depict proof-of-concept methods for potential future clinical applications including ACL injury risk screening, injury prevention training, and rehabilitation assessment. By synthesizing these results, we describe important opportunities that exist for using sophisticated modeling techniques to enable more accurate assessment along with standardization of data collection and creation of large benchmark datasets. If successful, these advances will enable widespread use of portable-sensing approaches to identify ACL injury risk factors, mitigate high-risk movements prior to injury, and optimize rehabilitation paradigms.

## 1. Introduction

Anterior cruciate ligament (ACL) injury is common in sports, with an estimated 400,000 people injuring their ACL in the United States each year [1], leading to over 129,000 ACL reconstruction (ACLR) surgeries [2]. Concerningly, nearly half of these patients are under 20 years of age, and they suffer from not only over 20% reinjury rates [115, 116] but also 50-80% knee osteoarthritis rates within a decade of injury [3, 4]. Knee osteoarthritis can lead to chronic pain and significant disability requiring surgical treatments such as total knee arthroplasty.

Preventing injury is a major goal of biomechanical assessments which seek to identify individuals at high risk of injury and provide feedback to prevent high-risk movement patterns. Identifying those with high injury risk and training them to adopt less risky movement patterns can lead to a wide range of health, societal, and economic benefits, including reductions in injury rates, sports drop-out rates, knee osteoarthritis incidence, and financial costs associated with rehabilitation and symptom management [5, 6].

After an ACL injury, the goal of biomechanical assessment is to guide and monitor the progress of comprehensive rehabilitation to allow return to sport and other physical activities without reinjury. Rehabilitation following ACLR should be customized according to various individual factors, like the amount of healing present in the ACL graft, activity level, and personal preferences [7]. Periodic biomechanical assessment can provide insights into patients’ recovery status and allow for customized physical rehabilitation protocols, thus accelerating recovery, lowering the risk of a secondary ACL injury, and helping athletes return to pre-injury sports level [8, 9].

Past research using laboratory-based biomechanical assessment has demonstrated that, for healthy athletes, non-contact ACL injury risk is associated with kinematic parameters such as the knee flexion and abduction angles (dynamic knee valgus), and kinetics parameters such as the knee extension and abduction moments during jump-landing and cutting [10-14]. Traditionally, measurement of these parameters requires optical motion capture and force plates. Although these devices are considered the gold standard for measurement, they confine the assessment to specialized motion laboratories, making evaluation inaccessible to a majority of people. Portable sensors such as inertial measurement units (IMUs) and smartphone cameras are less expensive and more portable, making them promising for out-of-lab assessment of pathologies like osteoarthritis [15], atrial fibrillation [16], and Parkinson’ s disease [17]. Similarly, these sensors may offer tremendous opportunities for less expensive, widespread ACL injury risk screening and injury-prevention training.

For patients undergoing rehabilitation after ACL injury and ACLR, kinematics and kinetics during squatting and walking may reveal dysfunctional movement patterns [18-20]. Although patients’ readiness to return to sport is traditionally assessed via strength and hop tests, recent studies suggest that readiness could be more holistically assessed based on kinematics and kinetics during running [21, 22], squatting [23, 24], and single-leg drop vertical jump [25]. However, tools that are ready for at-scale, clinical assessment of kinematics and kinetics do not yet exist. Recent portable-sensor-based assessment methods can be used in clinics or homes [26-28], thus increasing accessibility and affordability, and potentially benefiting thousands of patients following ACLR.

Accessibility of portable-sensor-based assessments can enable their broad use in ACL injury prevention and rehabilitation. However, the ideal methods to use, which clinically relevant parameters to assess, and at what point in the clinical workflow they should be employed remain unclear. We undertook this review to summarize the existing portable-sensor-based ACL assessment literature, including current target motions, sensing approaches, modeling techniques, and clinical applications. We also offer our perspectives on (1) future work that is necessary to achieve greater clinical impact and (2) new opportunities that may enhance the validity, reproducibility, and generalizability of the assessment methods.

## 2. Results

Our search yielded 1344 articles, of which 49 articles were included (Fig. 1), dating from 1990 to 2022. 98% of articles were published since 2007 and 51% since 2019 (Fig. 2a). IMUs were the most common sensor used in isolation (22%), followed by depth cameras (16%), red-green-blue (RGB) cameras (8%), and electromyography (EMG) (4%) (Fig. 2b). Kinematic parameters were the dominant target (71%), followed by spatiotemporal parameters (18%), kinetics (12%), and muscle activation (10%) (Fig. 2c). The sum of percentages is greater than 100% because several studies targeted parameters in multiple categories. Direct feature extraction (37%) was the most common analysis approach, followed by physics-based modeling (24%), and machine learning (22%) (Fig. 2d). The majority of studies used custom methods in their analysis (71%), while the remaining studies (29%) used direct outputs from commercially-available systems. ACL injury risk screening (57%) and rehabilitation assessment (55%) were the most common clinical applications, with a smaller percentage focusing on injury prevention training (6%). The sum of percentages is greater than 100% because several studies targeted multiple clinical applications.

**Fig. 1.**
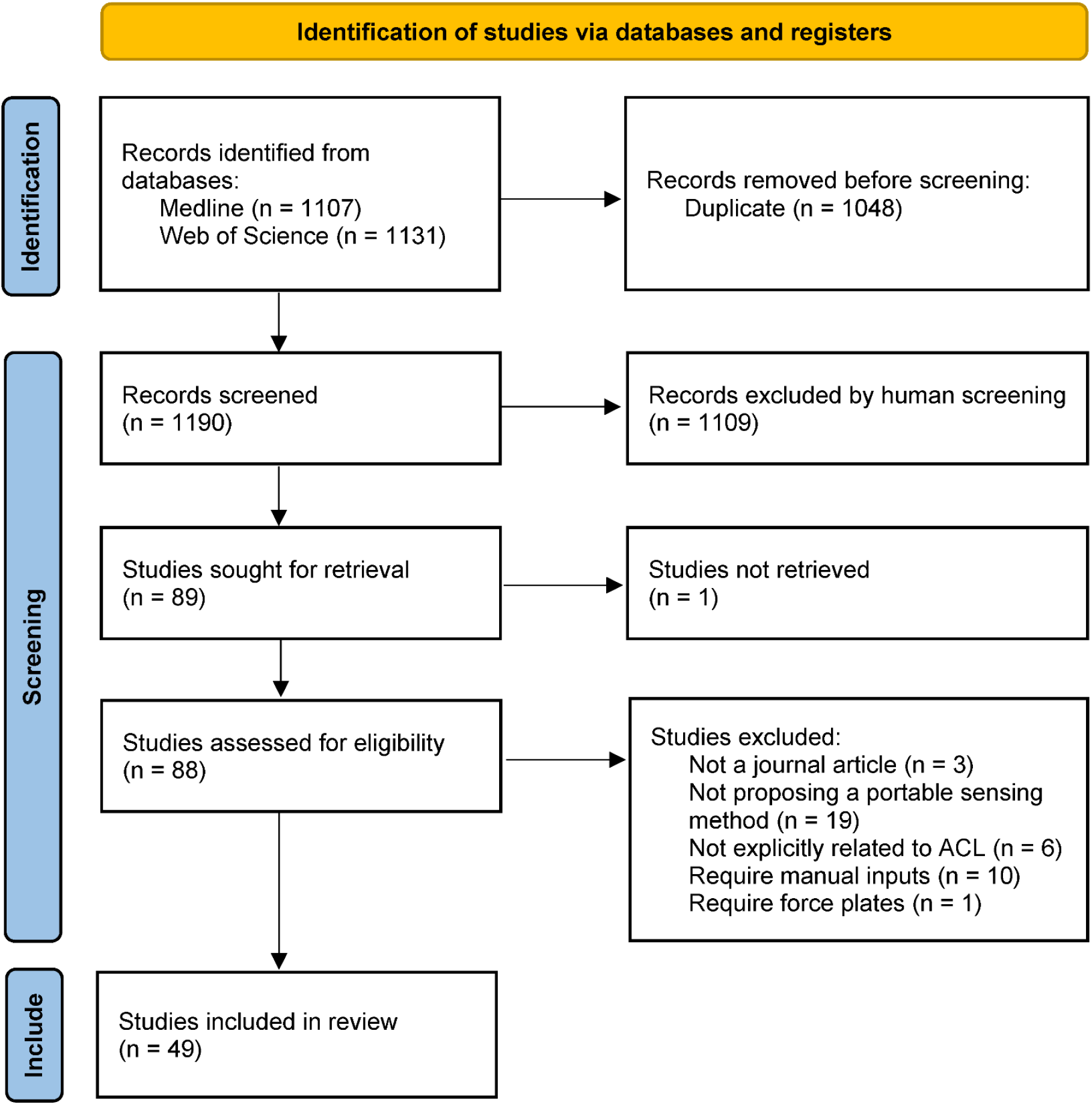
PRISMA flow chart. Search and study selection process for this review.

**Fig. 2.**
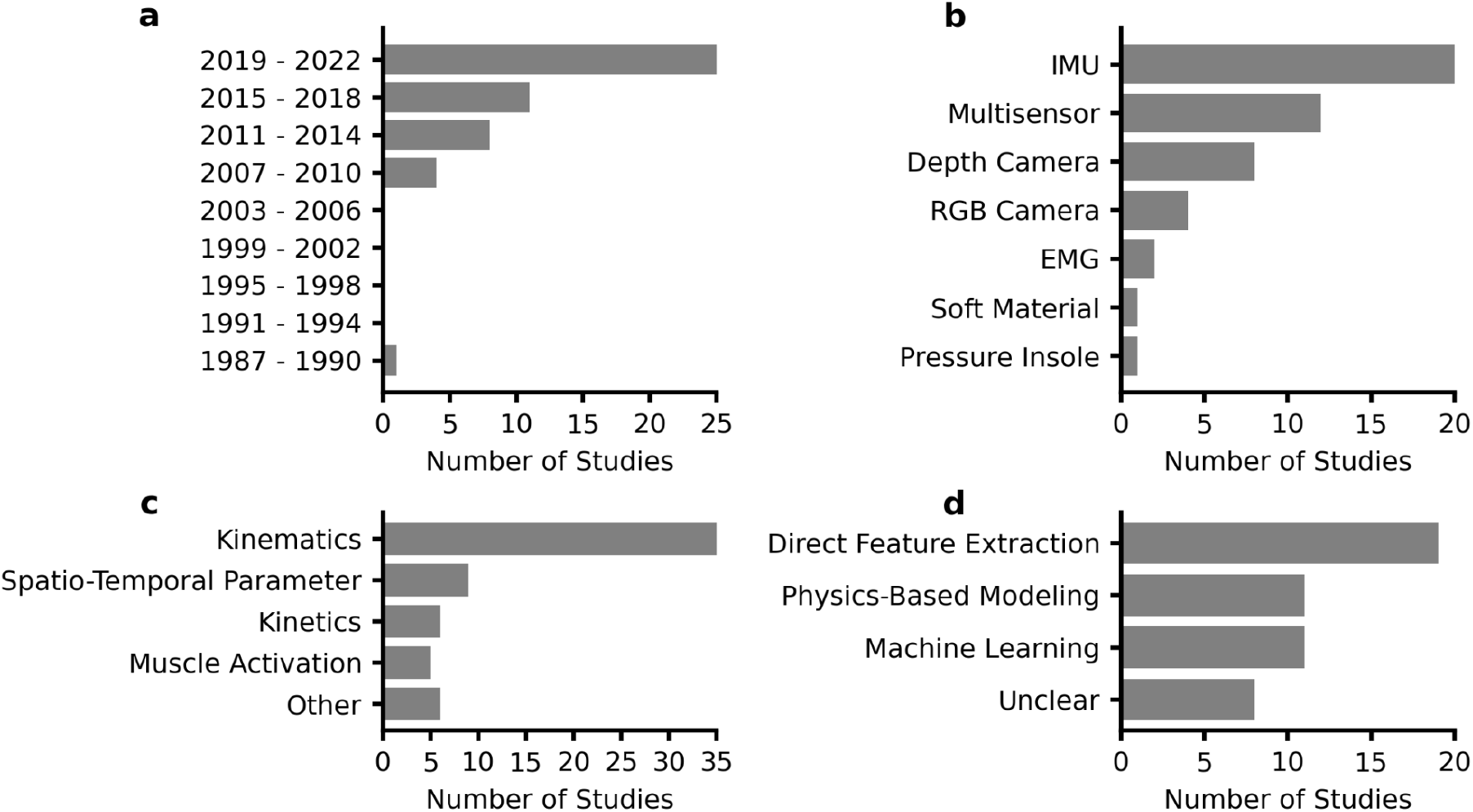
Characteristics of the included studies. **a** The number of articles has continually increased since 2007. **b** IMU was the most common sensor. “Multisensor” represents studies that used two or more sensing modalities. **c** Kinematic outcomes were most commonly estimated. All of the outcome measures used in each study were counted in the totals. **d** Direct feature extraction was the most common methodology.

### 2.1. Sensing Approach

IMU-based and RGB-camera-based studies had diverse configurations in terms of the number and placement of sensors (Table 1). The configuration of IMUs ranged from using one IMU for capturing shank movement to using seven IMUs for capturing all the lower-body joint kinematics. IMU sensors were most commonly placed on the shank, followed by the thigh, foot, and waist. The configuration of RGB cameras ranged from using one camera for capturing single-plane kinematics to using four calibrated cameras for capturing 3-D kinematics. The configurations of depth cameras were consistent in that seven out of eight studies placed a depth camera in front of the subject.

**Table 1.** Configurations of the included studies that only used one sensing modality.

| Sensor | Number | Placement | Paper |
| --- | --- | --- | --- |
| IMU | 1 | Shank | [29-31] |
| IMU | 1 | Waist | [32, 33] |
| IMU | 1 | Ear | [34] |
| IMU | 1 | Wrist | [35] |
| IMU | 2 | Thigh and shank | [36, 37] |
| IMU | 2 | Foot and shank | [38] |
| IMU | 2 | Both thighs | [39] |
| IMU | 2 | Both shanks | [40] |
| IMU | 3 or More | Multiple segments | [26, 41-48] |
| Depth camera | 1 | Frontal plane | [27, 49-54] |
| Depth camera | 1 | Sagittal plane | [55] |
| RGB camera | 1 | Frontal plane | [56] |
| RGB camera | 1 | Sagittal plane | [57] |
| RGB camera | 2 | Frontal and sagittal plane | [58] |
| RGB camera | 4 | Ceiling | [59] |
| EMG | 1 | Proximal to the patella | [60] |
| EMG | 3 | Vastus medialis and tibial tuberosity | [61] |
| Pressure insole | 2 | Foot | [62] |
| Soft fabric sensor | 1 | Above patella | [63] |

Included studies used six different types of multi-sensor combinations, and five of them involve IMU (Fig. 3). Seven studies designed algorithms to fuse multi-sensor data to estimate parameters [28, 64, 67-69, 73, 74], whereas the remaining five studies independently used different sensors to estimate different parameters [65, 66, 70-72]. Some sensor combinations might be redundant and could potentially be simplified. For example, four grounded optoelectronic bars were used to detect the initial foot-ground contact during landing alongside a shank IMU [73, 74]. The optoelectronic bars might be unnecessary because an IMU can estimate foot-ground collision by detecting the acceleration impulse [37]. Also, two studies simultaneously used a shank-worn IMU to measure tibial acceleration and a goniometer to measure knee flexion angle [70, 71]. The goniometer, which needs four belts to be strapped to the knee, could be replaced by an additional IMU on the thigh.

**Fig. 3.**
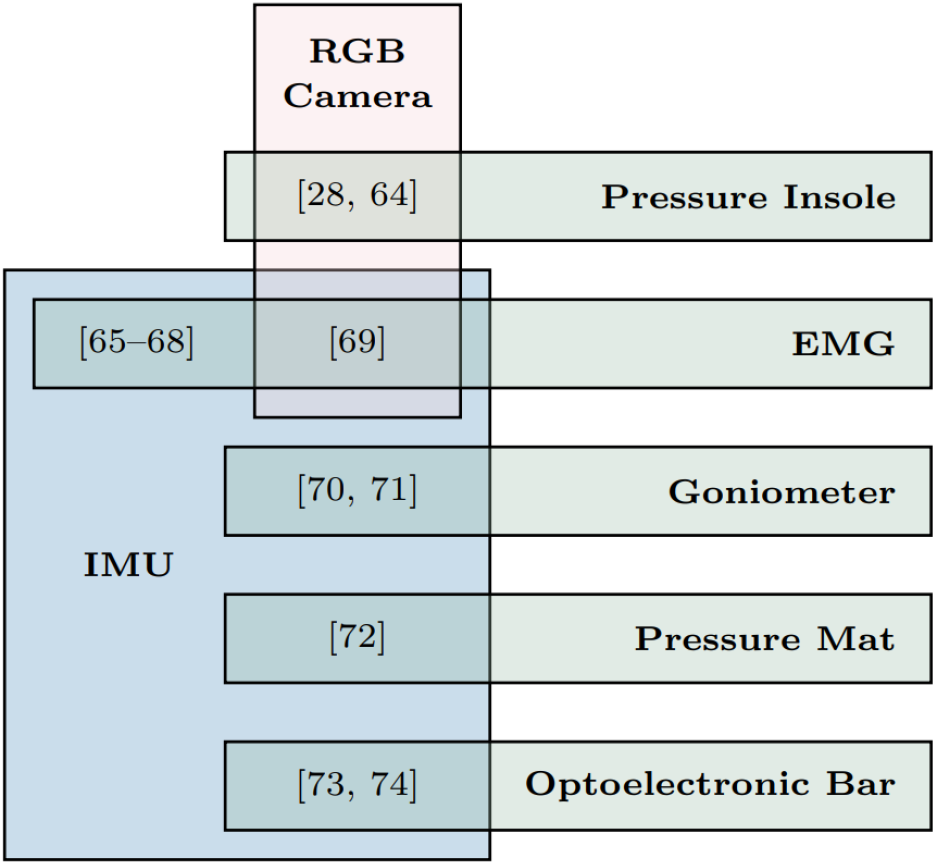
Studies used multiple sensing modalities. Eleven studies simultaneously used two sensing modalities and one study used three sensing modalities.

### 2.2. Target Motions and Biomechanical Parameters

Jump-landing, cutting, gait, and squatting were the most commonly measured activities in our included sample. Highlighted cells in Table 2 are primary parameters associated with ACL injury risk [10, 11] and rehabilitation outcomes [19, 20] that were identified in previous systematic reviews. Green cells are parameters that have been investigated using portable sensing, while red cells remain unstudied and represent important future opportunities. Many included studies estimated two primary kinematic parameters associated with ACL injury risk - knee flexion angle and abduction angle, mostly using IMUs, depth cameras, and RGB cameras (Table 2). Several studies estimated four primary parameters associated with ACL injury risk during jump-landing tasks, i.e., knee extension moment, knee separation distance, knee internal rotation, and vertical GRF during jump-landing tasks. Although knee abduction moment is a primary parameter that has been associated with ACL injury risk [10, 11] as well as knee osteoarthritis [77, 78], only one study estimated it during squatting [28]. Apart from highlighted primary parameters, other target parameters can provide insights into ACL injury risk and rehabilitation outcomes as well. For example, trunk kinematics during drop jump [118] and the “LESS” score (Landing Error Scoring System) [117] can be used to identify athletes with risky movement patterns. In addition, some “non-primary” target parameters are correlated with primary parameters, e.g., tibial acceleration with knee extension moment during drop landing (r = 0.72) [30], thigh angular velocity with knee extension moment during single-leg forward hopping (r = 0.59) [48], tibial angular velocity with knee extension moment during gait (r = 0.76) [40], and tibial and thigh angular velocity with knee abduction moment during drop vertical jump (r = 0.28-0.51) [43].

**Table 2.**
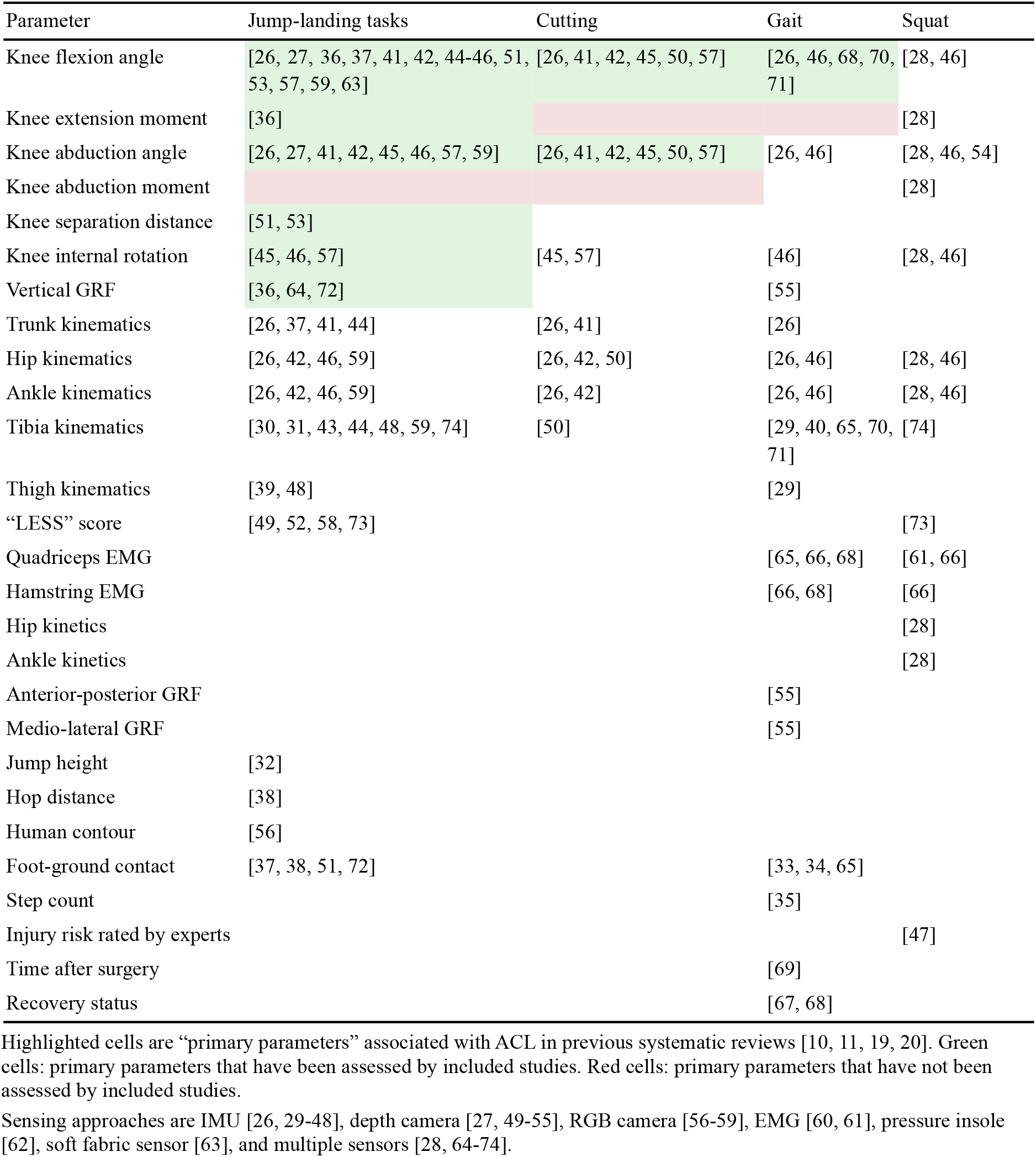
Target motions and biomechanical parameters of included studies.

Three studies conducted injury prevention training during drop vertical jumps by combining visual feedback with wearable IMUs [44], an RGB camera [56], or a depth camera [53]. In a drop vertical jump, the subject drops off a box, lands with both feet on the ground, and then immediately performs a maximum height vertical jump. The first study trained subjects to control their knee flexion angle and trunk lean estimated by three IMUs, and the training outcomes included increased knee flexion angle, increased trunk lean, reduced thigh angular velocity, and reduced knee abduction moment [44]. The second study trained subjects to maximize the overlap between their body contour estimated by an RGB camera and the contour of an expert movement, and the training outcomes included reduced vertical ground reaction force and ankle dorsiflexion moment [56]. The third study trained subjects to increase their knee separation distance estimated by a depth camera, and the training outcomes included increased knee flexion angle and knee separation distance [53]. Although these training regimes have been demonstrated effective in modifying ACL injury risk factors, their utility in reducing real-world injury incidence rates have not been prospectively validated.

### 2.3. Accuracy and Reliability

The validity of the sensing approaches proposed by 22 studies (45%) was examined against the gold standard from force plates, optical motion capture, and human raters. Three studies (6%) examined their validity against parameters measured by another portable sensing system, i.e. knee angles from goniometers [63], knee angles from a commercial IMU system [46], and step time asymmetry from pressure insoles [34]. The remaining 24 studies (49%) did not examine the validity of the estimated parameters. There were substantial differences in the accuracy metrics used across studies, making it challenging to compare the performance of different approaches. IMU-based studies reported root mean square errors (RMSEs) of 1.1 - 6.5 deg for knee flexion angle estimation [41, 26, 37, 45] and 3.3 - 10.9 deg for knee abduction angle estimation [41, 26, 45]. The accuracy of knee abduction angle estimation was poor considering the small knee abduction range of motion. The RMSE of knee flexion angle estimation was 6.8 deg when using eight calibrated RGB cameras [28], while the RMSE was as low as 1.7 deg when using one single RGB camera and two reflective boards with Moiré patterns attached to the thigh and shank [57]. Three studies examined the reliability of the sensing approach, either within-day [33, 63] or between-day [74] test-retest repeatability based on intraclass correlation coefficients (ICC). Excellent repeatability was observed in the knee flexion angle estimated by a soft fabric sensor (ICC ≥ 0.9) [63].

### 2.4. Methodology of Biomechanical Parameter Estimation

We categorized the methodologies for the analysis of the acquired data into three separate categories: 1) physics-based modeling that includes studies with kinematics reconstructed from raw sensor measurements and kinetics estimated via inverse dynamics or musculoskeletal models, 2) machine learning models to estimate subjects’ status or estimate parameters, and 3) direct feature extraction using investigator-defined parameters from the raw sensor data.

Eleven studies primarily used physics-based modeling. Integration of gyroscope data was combined with several drift compensation methods to estimate the sensor and body segment orientation in eight investigations [37, 38, 41, 44-46, 68, 74], and seven of these studies used the relative orientation between two segments to derive joint angles [37, 41, 44-46, 68, 74]. Only one study used musculoskeletal modeling, which estimated the GRF by simulating 25 artificial muscle-like actuators placed under each foot [55].

Most of the machine learning studies focused on building classification models, whereas only one study built a regression model (linear regression) to predict kinetic parameters [36]. Two other studies implemented an existing deep-learning-based keypoint detection algorithm, OpenPose [119], to estimate joint centers from 2-D videos; one of these studies used the keypoints to derive planar joint angles [58] and the other predicted the “LESS” score [59]. Classification models including Support Vector Machines [47, 65, 73], Linear Multinomial Logistic Regression [47], Decision Trees [47, 73], Naive Bayes [47], K Nearest Neighbors [47, 73], and Fuzzy Clustering [67, 69] were used to identify walking states, identify subjects at high ACL injury risk, and classify the time (e.g., 0-3 months, 3-6 months, and 6 months or more) after ACLR surgeries.

A few studies directly used raw sensor measurements for assessment [30, 31, 33, 34, 39, 40, 43, 48, 61-64, 70-72], for example, extracting peak acceleration and angular velocity from the thigh or shank IMU data for assessment [30, 31, 39, 40, 43, 48, 64]. Characteristics of the IMU data (e.g., local peaks) can also be used to segment gait cycles and derive step time asymmetry [33, 34].

### 2.5. Experimental Design

The number of subjects recruited in the included studies ranged from 9 to 169, with the median being 24 (Fig. 4a). Twenty-five studies (51%) did not exhibit biases across the sex of included subjects, in that the percentages of females were between 34% and 66% (Fig. 4b). Importantly, five studies (10%) focused on females [35, 36, 53, 70, 71, 73, 74] (Fig. 4b), as females are more than twice as likely as males to have a first-time non-contact ACL injury [80]. However, four studies (8%) recruited only male subjects without providing a scientific rationale. Twenty-one studies (43%) recruited patients following ACLR. The time from ACLR to experimental testing were within 3 months [33, 34, 40, 60, 61, 65], 3-12 months [28, 29, 32, 34, 39, 40, 46, 48, 62, 64, 65, 67-69, 72], or more than 12 months [35, 38, 48, 72] for the recruited patients. Eight studies (16%) recruited athletes at high risk of ACL injury, including basketball players [73, 74], soccer players [42, 59], and gymnasts [53]. Eight studies (16%) explicitly reported that their experiments were performed out of lab, including clinics [34, 46, 64], hospitals [33], soccer fields [42], and unconstrained daily life [35, 62, 65].

**Fig. 4.**
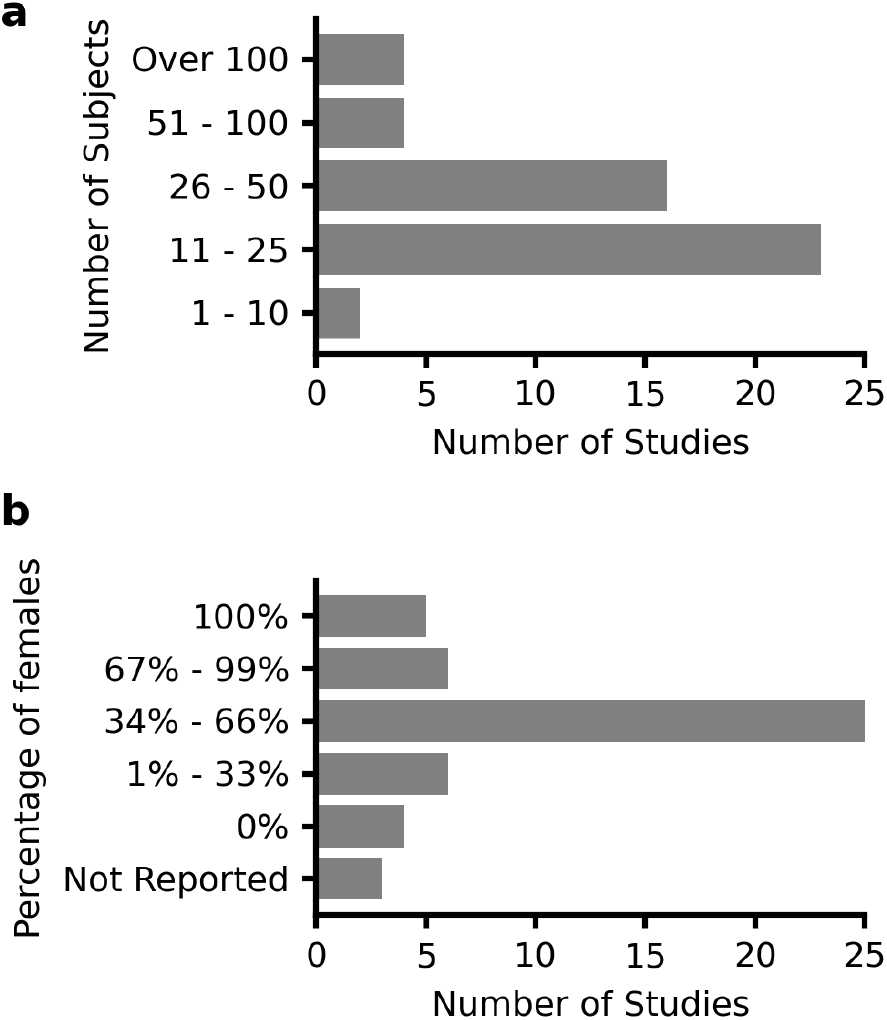
**a** Number of subjects and **b** percentage of female subjects.

None of the studies that estimated ACL injury risk factors prospectively evaluated their estimation results against subjects’ future injury occurrence. One study recruited thirteen basketball players, and one of them suffered from an ACL injury within two weeks after the first test session [74]. The injury was caused by an incorrect landing phase after a single-leg jump with a pivot-shift mechanism. Analysis of shank-worn IMU data revealed that the variances of her tibial orientation and acceleration were significantly larger than the 12 uninjured players during a countermovement jump test, indicating poor leg stability and load absorption capability. For rehabilitation assessment, six studies (12%) have used portable sensors to track post-surgery longitudinal changes of quadriceps EMG [60, 61], step count [35, 65], and level of gait asymmetry [34, 35].

### 2.6. Readiness for Deployment

We categorized the included studies into three stages based on their readiness for deployment (Fig. 5). 72% of the included studies are in stage I, development and prototyping, as they proposed a novel method in-laboratory and associated its outcome with ACL injury risk or rehabilitation status. 24% of the included studies are in stage II, preliminary clinical validation, as they demonstrated their clinical utility. These studies successfully used portable sensors to detect deficits in patients following ACLR [62, 64, 65, 70, 72], identify subjects with high-risk movement patterns [37, 42, 47, 52, 73], and enable injury prevention training that led to significant reductions in ACL injury risk factors [44, 53, 56]. However, before the deployment of these studies, further clinical validation is needed to validate their test-retest reliability as well as effectiveness in accelerating the post-ACLR recovery, or predicting and reducing real-world injury incidence rate. 4% of the included studies are in stage III, clinical validation, as they have proved their utility among the target population (e.g., athletes, patients following ACLR) outside of laboratories (e.g., sports fields, clinics). The only two studies in this stage monitored the longitudinal changes of quadriceps EMG and performed feedback training during rehabilitation exercises among an ACLR cohort [60, 61], and one of them was published in 1990 [60]. Randomized controlled trials showed that EMG biofeedback accelerated the recovery of quadriceps strength and knee range of motion in early-stage rehabilitation.

**Fig. 5.**
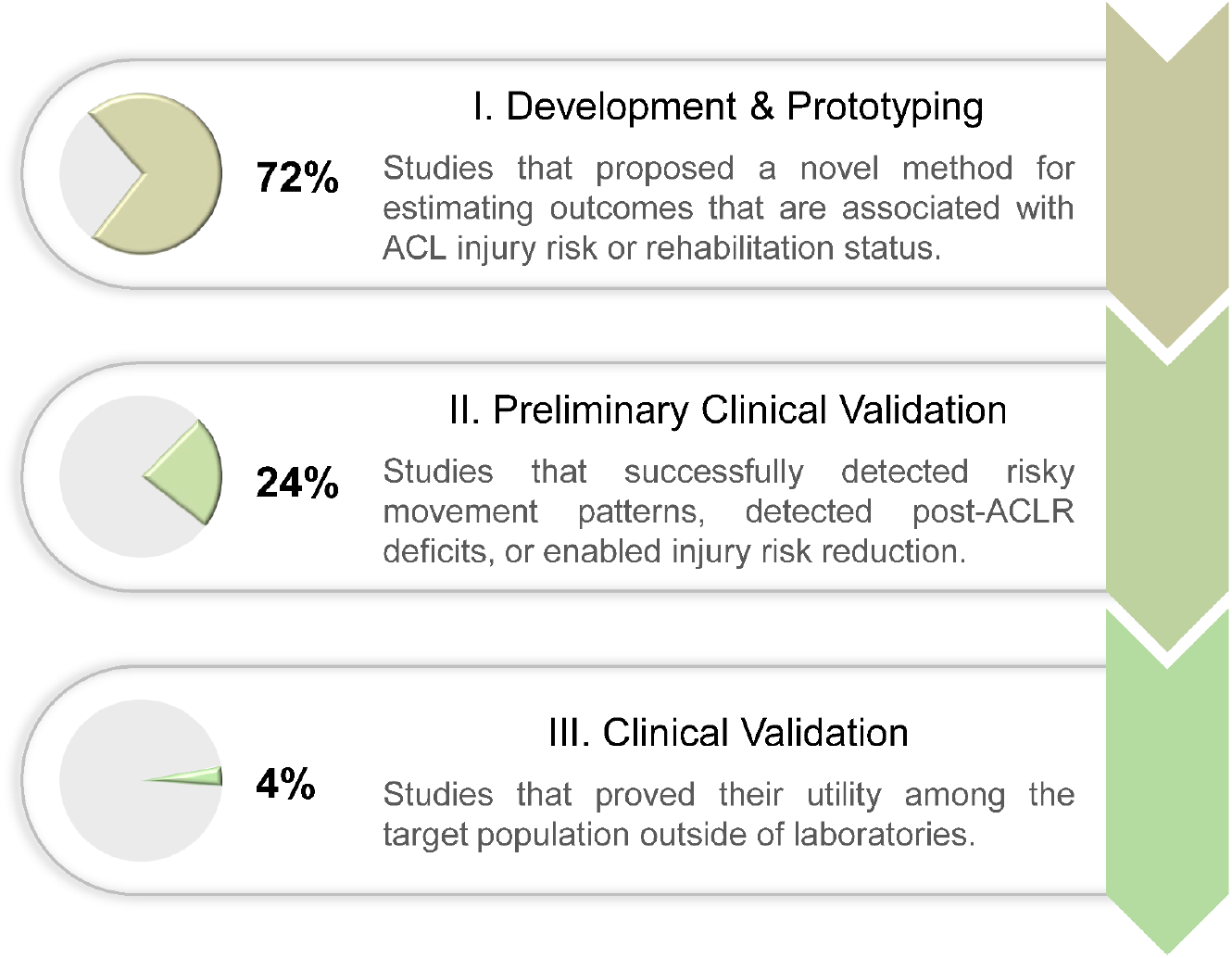
Categorization of the included studies into three stages based on their readiness for deployment.

## 3. Discussion

The use of portable sensing for ACL injury risk stratification and rehabilitation has been increasing at an accelerated rate, with 98% of our reported studies occurring since 2007 and 51% since 2019 (Fig. 2). These studies have been dominated by the use of IMUs (43%) as well as depth and RGB cameras (24%). Such portable technologies for assessing mechanics have primarily been leveraged to collect kinematic data at the knee during jump-landing tasks, and, to a lesser extent, during cutting and gait motions. While these studies have laid important groundwork for portable sensing of ACL injury and rehabilitation, this review highlights important strengths, current knowledge gaps, and future opportunities for the underlying technologies as well as their clinical applications. In the following, we provide our perspective on how to improve existing modeling methods and achieve broader clinical impact.

### 3.1. Clinical Applications

#### 3.1.1. Injury Risk Screening

Using portable sensing to estimate traditional parameters or identify new parameters associated with ACL injury risk represents a significant research opportunity. Jump-landing tasks were the most popular dynamic motion among included studies and have been established as being well suited for identifying athletes at greater risk of ACL injury [10]. Cutting motions were less popular, but could be complementary to jump-landing tasks because knee kinematics and kinetics were significantly different during these two motions when assess in-lab [14, 79]. Thus, simultaneous evaluation of jump-landing and cutting movement quality would provide a broader array of real-world conditions for more reliable injury risk screening. Systematic reviews of lab-based assessments have established that the knee angles, knee moments, and vertical GRF during jump-landing tasks and cutting are primary parameters for understanding ACL injuries and rehabilitation [10, 11]. Among these primary parameters, knee angles were estimated by many of the reviewed studies; however, only one included study estimated knee extension moment [36] and no study estimated knee abduction moment during jump-landing tasks or cutting. Recent research has shown that knee abduction moment during gait can be estimated from simulated 2-D video data using neural networks [81] or from real 2-D video data using neural networks and musculoskeletal simulation [82]. Future research should test these methods for dynamic activities relevant to ACL injury risk screening, such as jump-landing and cutting motions.

A particular challenge with studies to establish new metrics for injury risk screening, however, is the low occurrence of ACL injuries that makes it challenging to pair pre-injury mechanical patterns to injury occurrence. Previous studies recruited hundreds to thousands of athletes from high-risk sports (typically young female basketball and football players) to validate traditional ACL injury risk factors such as joint laxity measured using a knee arthrometer [83] and knee abduction moment measured by marker-based motion capture and force plates [13]. In contrast, despite the portable nature of the sensing approaches, the included studies estimated parameters with 9-169 subjects (the median is 24), and only 16% recruited athletes. Consequently, ACL injury incidence was rare among the included studies (only one study reported an incident [74]), suggesting the need for future studies that validate clinical utility through prospective evaluation of the estimated parameters against real injury occurrence. One solution to scarcity of data from injured athletes is the creation of a standardized pipeline of sensor deployment, data processing, archiving, and data sharing, which could enable multi-center data collection from a large number of athletes in the real world. Deployment of such a multi-center effort is particularly feasible for non-wearable sensors (e.g., RGB and depth cameras), which can passively collect data without the burden of donning and doffing the sensors. Furthermore, such datasets will enable the identification and prospective validation of novel parameters that may more accurately predict ACL injury risk than traditional parameters.

#### 3.1.2. Injury Prevention Training

The prospective studies of ACL injury risk mentioned previously will be pivotal for providing targets for preventative training. It has been shown that young athletes can be trained to adopt less risky posture and reduce injury risk [12]. However, effective training typically requires multiple sessions per week during both pre-season and in-season [84] along with verbal feedback from expert coaching staff based on subjective visual observations [85]. In reality, with practical constraints such as demanding exercise protocols for athletes and limited availability of coaches knowledgeable in injury prevention, such training and feedback are challenging to implement. Prior studies have demonstrated that the subjectivity of such feedback may also lead to athletes experiencing considerable variability between recommendations amongst coaches [86]. In contrast to the subjective verbal feedback from human observations, the visual feedback enabled by portable sensors is tangible, quantifiable, and more importantly, objective [44, 53, 56]. Further, the capabilities for automatic tracking of training and progress provided by portable sensing technologies could also open the door to new strategies, such as gamification, to motivate and engage athletes in completing injury prevention programs [121, 122].

Current efforts focus on the paradigm described above for testing specific movements, providing feedback, and athletes learning to change that movement. However, lab-based methods cannot measure risk factors during practice or game-play; thus real-world changes to risky behaviors in response to training are unknown. Portable sensors are now commonly used to measure athletes’ position in field sports using GPS, or movements using IMUs [87], from which ACL injury risk factors can be extracted. Longitudinal tracking of risk factor improvements during practice and game-play provides a novel avenue to determine the effectiveness of injury prevention training. Additionally, in-game and in-practice tracking allows data collection in a much larger volume in terms of the duration and the number of athletes compared to traditional in-lab environments. The sheer scale of data may overcome relatively lower quality of portable sensors and enable the identification of novel risk factors that were not possible in a solely lab-based environment.

#### 3.1.3. Return-to-Sport Decision Making

Patients following ACLR need 4 - 12 months of rehabilitation to restore movement quality [88, 89]. After rehabilitation, patients’ readiness to return to sport is commonly determined by whether their inter-limb symmetry in muscle strength, hop distance, and hop task completion time are larger than 90% [75, 76, 90]. While these tests are beneficial since they can be implemented in clinics, previous studies reported that strength tests lack functional relevance to sporting situations [91]. Furthermore, it was reported that since hop tests only indirectly assess knee function and loading, they may mask asymmetry in lower limb biomechanics [90, 92-94]. Hop tests are also sensitive to small alterations in the test procedures [95].

Apart from strength and hop test measures, asymmetrical knee kinematics and kinetics during gait and double-leg squat as well as improper single-leg squat mechanics such as increased knee abduction angle may reveal dysfunctional movement patterns [19, 20, 96]. One barrier impeding the adoption of parameters during squatting and gait into return-to-sport decision-making is their reliance on force plates and marker-based motion capture from specialized laboratories. Many studies have proposed portable-sensing methods to estimate knee kinematics (Table 2). Also, as previously mentioned, portable sensors coupled with neural networks and musculoskeletal simulation have shown promise at predicting knee kinetics [36, 81, 82]. Using these technologies to develop assessments that are fast and accurate enough for clinical deployment could enable better return-to-sport decision making and potentially lower the risk of reinjury. Clinical validation studies could start with young athletes [115, 116], since their high reinjury rate could increase statistical power. Additionally, the ability to track biomechanical changes over time will both inform the rehabilitation approaches of clinicians and promote long-term patient engagement.

### 3.3. Modeling Technique

Most of the included studies used direct feature extraction and IMU data integration with drift compensation to estimate parameters; however, they cannot provide insights into GRF or joint kinetics. In contrast, machine learning, despite being computationally expensive and prone to overfitting in low-data regimes [97], has shown its potential for estimating kinetics in an end-to-end manner [98-100] or in combination with musculoskeletal modeling [82]. Few studies have attempted to use machine learning to estimate kinetic parameters associated with an ACL injury, so this field remains unstudied and represents important future opportunities; here we offer a few ideas.

- Using transfer learning to augment training data for machine learning models in conditions with limited data. For example, machine learning models can be pre-trained on large corpora of data collected during easier-to-measure motions (e.g., walking) and/or data synthesized from readily available sources (e.g., [101]). Subsequently, those models can be fine-tuned on data collected during drop vertical jump or cutting where massive data collection is more difficult.
- Incorporation of machine learning to enhance physics-based modeling. For example, physics-based models simplify the human body using parameterized formulas, where parameters were traditionally determined empirically or using population averages. Alternatively, these parameters can be learned from collected data using machine learning [102, 103].
- Extraction of 3-D joint angles from camera data. Calculation of 3D joint kinematics requires collection of ≥ 3 non-collinear markers per rigid segment. Body keypoint detection algorithms employed by the included studies only extract joint centers, making derivation of 3-D joint angles (flexion, abduction, and internal rotation) impossible. Future studies may consider employing novel algorithms for extracting body meshes [104-107] or conventional biomechanical markersets that can be used to derive 3-D angles.

Machine learning models have a strong dependency on datasets, so it can be challenging to generalize the results to new datasets. To prevent overfitting and guarantee the models’ reliability and generalizability, we suggest future studies follow the general recommendations provided to biomechanists by Halilaj and colleagues [120]. Here we offer a few additional recommendations for ACL-related assessment.

- Use the target population for model training and testing. If the dataset is from a general population cohort, the model may not generalize to an athlete cohort who are stronger and faster, or to a patient cohort with pathological movement patterns.
- Use a large and representative dataset, for example, a dataset collected from multiple laboratories with a balanced distribution in sex, height, weight, and age of included subjects. A small dataset collected by one single operator using one set of devices can suffer from biases such as biased marker positioning [108]. If the model is trained on such datasets, whether those biases are inherited by the model needs to be investigated.
- Do not test the model using the data from the subjects that were involved in model training, unless additionally training data can be easily acquired from new subjects in practical applications.

### 3.4. Dataset and Benchmarking

Benchmark datasets are curated, publicly available sets of data for enabling objective comparisons between studies and rigorous selection of state-of-the-art methods [109]. They have already proven their fundamental importance in research areas such as computer vision. Although many included studies estimated the same biomechanical parameter during the same type of motion (Table 2), significant differences exist in their recruited cohorts of subjects, sources of ground truth, and metrics of validity. It is therefore difficult to impossible to truly compare the validity, sensitivity to parameter changes, or test-retest repeatability between these methods. Although a few comprehensive biomechanical datasets containing multiple portable sensors have been published [110-112], none have focused on ACL-specific tasks. To make it easier for future researchers to select the optimal sensing approach and estimation method, we here call for collection and publication of a benchmark dataset, ideally following a standardized pipeline (section 3.1.1) and open science principles to ensure its findability, accessibility, interoperability, and reusability [113]. Such an ideal dataset should contain simultaneously collected ground-truth kinematics, GRF, and multiple portable sensor data (Table 1 and Fig. 3) during a range of ACL-specific motions (Table 2). A dataset that includes repeated measures on the same or separate days will enable assessment of measurement reliability, and thus calculation of the minimum detectable change which is crucial for clinical use on an individual basis [114].

## 4. Conclusion

In this manuscript, we summarize the state of using portable sensors to enable a range of ACL-related biomechanical assessments. Through these studies, we showed that portable sensing can be used to monitor patient progress through the rehabilitation process and train athletes to reduce injury risk factors. However, despite their promising results, we highlight numerous opportunities that exist in clinical validation of the methods proposed by the reviewed portable sensor studies. We also highlight two important opportunities for future research in: (1) exploring sophisticated modeling techniques to enable more accurate assessment and (2) standardizing data collection and processing methods to pave the way for procurement of large benchmark datasets, and multi-center trials for clinical validation. The sheer amount of portable sensor data may enable large-scale prospective studies for the identification of new ACL injury risk factors, leading to novel targets for preventative training. The capabilities for automatic tracking of training and progress provided by portable sensing technologies could open the door to new strategies, such as gamified platforms, to motivate and engage athletes and patients in completing training programs. If successful, these advances will enable widespread use of portable-sensing approaches to estimate ACL injury risk factors, mitigate high-risk movements, customize rehabilitation paradigms for improved long-term health outcomes, and quantify return-to-sport readiness.

## 5. Methods

## 5.1. Literature Search Approach

The systematic review was conducted in accordance with the Preferred Reporting Items for Systematic Reviews and Meta-Analyses (PRISMA-2020) guidance [123]. We searched articles published up to March 6, 2022 from the following databases: Medline (1950-) and Web of Science Core Collection (1950-). The search focused on retrieving articles that included: (1) ACL, (2) portable sensing approaches such as IMU, EMG, video, and pressure insole, and (3) clinical applications such as injury risk screening (Table 3). We only considered articles written in English and those contained at least one term from each of the three categories above either in their title, abstract, or keywords.

**Table 3.**
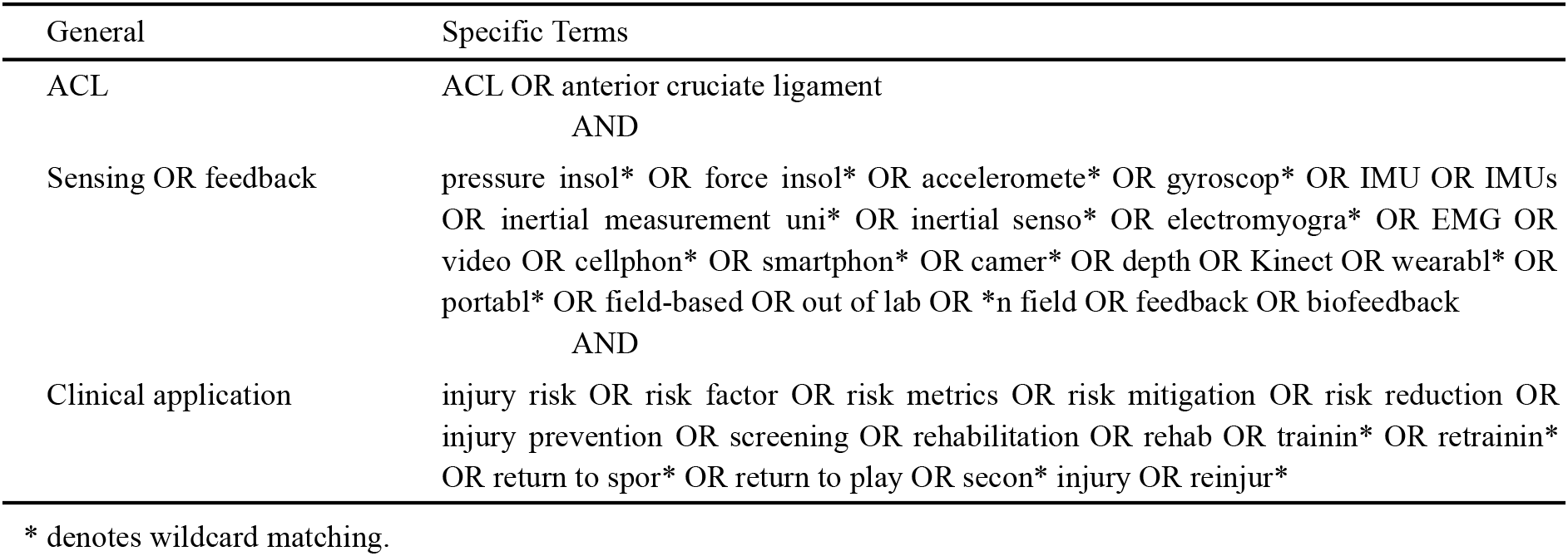
Specific search terms used for the literature review

### 5.2. Inclusion and Exclusion Criteria

Two authors (T.T. and A.A.G.) independently reviewed titles, abstracts, and keywords of all the retrieved articles. Inclusion/exclusion disagreements were resolved by full-text review and discussion to reach consensus. We excluded dissertations, theses, conference proceedings, and conference abstracts. We also excluded articles whose primary purpose was not development, validation, or use of portable sensing for ACL-related assessment. In addition, we excluded articles that did not involve human subjects, articles that required human raters or manual labeling for qualitative assessment, and articles that used force plates and marker-based motion capture measurements as input data for assessment. Articles were not excluded if those measurements were used as the gold standard to determine the validity of portable sensing approaches.

### 5.3. Outcome Extraction

We carefully read and extracted the following outcomes from the included articles: area of application (i.e., injury risk screening, injury prevention training, rehabilitation assessment), sensing approach (e.g., IMU, RGB camera, EMG), target motion (i.e., jump-landing tasks, cutting, gait, squatting), target biomechanical parameter (e.g., knee flexion angle, vertical GRF), category of the parameter (e.g., kinematics, kinetics), methodology of estimation (i.e., physics-based modeling, machine learning, direct feature extraction), developer of the method (i.e., academic laboratories, commercial companies), number of subjects, sex ratio, involvement of athletes, experiment site (e.g., laboratory, clinics), validation (e.g., validated against force plates and optical motion capture), and repeatability (e.g., test-retest reliability). We also assessed whether included articles validated their clinical utility in detecting patients’ recovery status, accelerating rehabilitation, predicting future ACL injury occurrences, or enabling feedback training for injury risk reduction. Some terminologies were unified or simplified if they depicted the same fundamental measurement. For example, knee flexion angle, knee extension angle, and sagittal plane knee angle were unified as knee flexion angle.

## Data Availability

All data produced in the present work are contained in the manuscript

## Author Contribution

A.S.C. and P.B.S. oversaw the development of the review. K.G.S., S.L.S, S.D.U, J.L.H., and S.L.D. conceptualized the study. T.T. and A.A.G conducted the literature search. T.T., A.A.G, and B.F. prepared the first draft of the manuscript. All authors participated in drafting the manuscript, critically reviewed drafts of the manuscript, and approved the final version to be submitted.

